# A Comparison of Two Deep Learning Approaches to Distinguish Functional Dissociative from Epileptic Seizures Using Event Videos

**DOI:** 10.64898/2025.12.18.25342568

**Authors:** Asala N Erekat, Mark Dakov, Jacky Cheung, Megan Mackenzie, Stefano Malerba, Ilana Lefkovitz, Andy Ho Wing Chan, Jiyoon Hwang, Felix Richter, Alec Gleason, Lara Marcuse, Madeline Fields, Benjamin S Glicksberg, Girish N Nadkarni, Nathalie Jetté, Benjamin R Kummer

## Abstract

**Background:** Differentiating between motor functional dissociative seizures (FDS) and motor epileptic seizures (ES) is a common diagnostic challenge, requiring video electroencephalography (vEEG) as gold standard. However, vEEG requires specialized technicians and clinical experts to set up and interpret and oftentimes fails to capture events. We sought to develop machine-learning (ML) tools to carry out this diagnostic task independently of vEEG or human review by a neurologist.

**Methods:** In this retrospective study, we developed two proof-of-concept ML models to differentiate motor ES from FDS based on video of FDS and ES events in patients who underwent inpatient vEEG monitoring at an academic medical center between 2012 and 2021. The first model employed a pose-estimation approach, using body landmark features that were labeled frame-by-frame by three neurologists. The second utilized an end-to-end 3D convolutional neural network (CNN), thereby learning directly from raw video frames. Using board-certified epileptologist review as a clinical gold standard, we measured model performance by area under the receiver-operating (AUROC) and precision-recall (AUPRC) curves, sensitivity, precision, and accuracy against a held-out test set of videos.

**Findings:** We included 101 unique patients with 106 total event videos, comprising 61 (60.4%) ES and 45 (44.6%) FDS events. Both ML models distinguished both seizure types better than chance. The pose-estimation-based model achieved an AUROC of 0.71, AUPRC 0.53, sensitivity 0.90, precision 0.50, and accuracy 0.62. The CNN model exhibited superior overall performance, achieving an AUROC 0.78, AUPRC 0.84, balanced sensitivity and precision (both 0.82), and accuracy 0.80.

**Interpretation:** Our findings demonstrate the superiority of CNN over pose-estimation models to differentiate between motor ES and FDS using video alone. Although future studies are needed, these models hold potential as adjunct diagnostic tools by enabling rapid, objective seizure evaluations without immediate neurologist involvement.

**Funding:** CTSA grant UL1TR004419 (Kummer), NIH grants R01DK133539 and R01HL167050 (Nadkarni), NIMH T32 grant MH122394 and AAN grant 19-1120 (Chan).

## INTRODUCTION

Differentiating epileptic seizures (ES) from functional dissociative seizures (FDS, psychogenic non-epileptic seizures, or PNES) is challenging for clinicians and is a principal cause of the average 7-year delay in diagnosing FDS^1,2^. While epilepsy is treated with anti-seizure medications and adjuvant surgeries, FDS is treated with psychotherapy.^3^ These treatment differences mean that misclassifications can result in significant medical risks and costs. These are compounded in status epilepticus (SE), which requires emergent care not indicated in FDS. Conversely, if SE is mistaken for FDS, treatment delays can lead to brain injury, respiratory compromise, or death.

The gold standard for differentiating ES from FDS is video electroencephalogram (vEEG), which is typically performed in specialized labs with dedicated staff and equipment. Access to EEG can be limited by socioeconomic factors, target events are frequently not captured^4^, and interpretation requires neurologists, who are scarce in supply worldwide. These bottlenecks have created a need for diagnostic alternatives to vEEG^5^.

With the ubiquity of smartphone cameras, videos of seizure events are increasingly commonplace. Although human review of videos (without EEG) can differentiate FDS and ES^5–11^, accuracy is highest among neurologists and lowest among non-neurologists.^12,13^ Due to the scarcity of neurological expertise, automated video analysis that obviates the need for human review is an attractive alternative to human video review. Two machine learning (ML) approaches have been used for seizure detection^14,15^: (a) pose-estimation models, which identify and track anatomical landmarks frame-by-frame as predictive features, and (b) deep-learning models such as 3D convolutional neural nets (3D-CNN), which directly learn spatiotemporal patterns from raw video.

Using both ML approaches, we developed two models to differentiate motor FDS and ES using only video (Figure 1). We hypothesized that both approaches were equivalent in performance. We exclusively focused on events with motor manifestations because these are readily detectable and clinically common.

**Figure 1.**
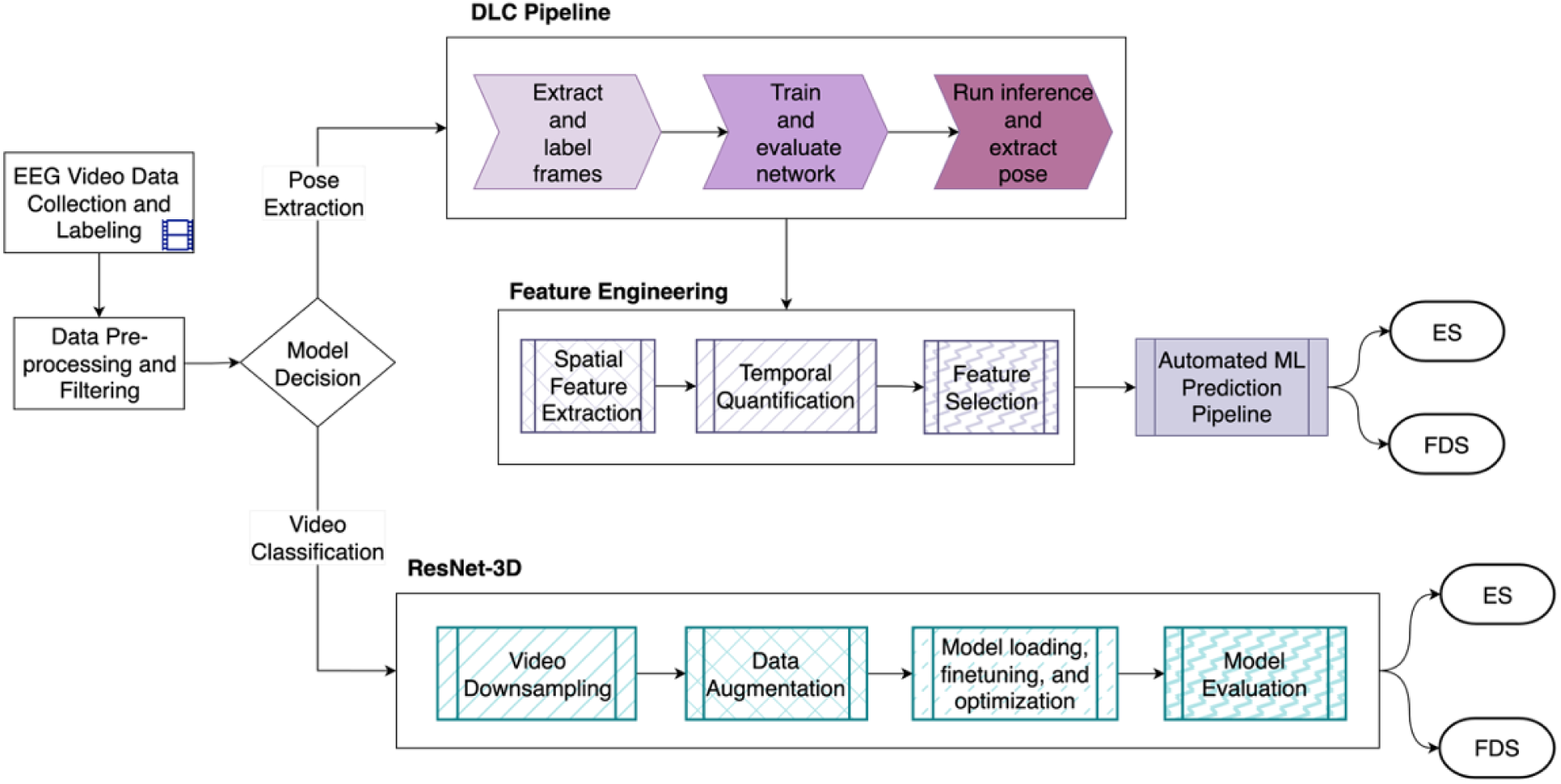
High-level overview of data processing pipeline encompassing both seizure classification approaches. Abbreviations: DLC, DeepLabCut; ES, epileptic seizure; FDS, functional dissociative seizure; ML, machine learning.

## METHODS

### Patient Population

We conducted a retrospective cohort study using data from patients 12-years and older who underwent inpatient vEEG between January 2012 and June 2021 at Mount Sinai Health System, a health network in New York City comprised of 7 clinical sites and affiliated with the Icahn School of Medicine at Mount Sinai (ISMMS). Each vEEG study was interpreted by a board-certified epileptologist in an EEG report, which was extracted from our institutional electronic health record (EHR). The ISMMS IRB approved the use of this patient data for research under protocol number 19-00738 and waived the requirement for informed consent. Because of challenges in de-identifying patient videos, data underpinning this study will not be made available, although model code is publicly available at https://github.com/asalaerekat/epilepsy-project.

A research coordinator (MD) manually reviewed all available inpatient vEEG reports on a shared hospital network drive to identify events with motor manifestations. Each event was categorized into ‘motor ES’ (generalized tonic-clonic, tonic, myoclonic-tonic-clonic, and focal to bilateral tonic-clonic), ‘FDS with motor manifestation’, or ‘other’ (e.g., movement disorders, convulsive syncope, other epileptic seizures including atonic, focal non-motor, myoclonic, etc). Events with exclusively non-motor symptoms or subtle motor findings unlikely to be captured by human review (e.g., eye fluttering) were excluded. We excluded inpatient vEEGs that lacked video data and any outpatient EEG studies. For all included patients, we extracted age, sex, race, any available pre-EEG brain MRI findings, and International League of Epilepsy categorization of epilepsy^16^ from the EHR.

### Video Processing

Videos were exported using our institutional EEG monitoring software (NeuroWorks, Natus Medical, Middleton, WI, USA). We used EEG report timestamps to identify the exact timeframe of each clinical event and exported events using Microsoft Windows Video Editor (Microsoft Corp., Bellevue, WA, USA). We excluded any video that had (1) excessive pixelation compromising visibility, (2) camera zoom levels that occluded full-body visibility, or (3) camera obstruction by an object or another person for more than 50% of the video. To ensure unbiased evaluation, we used a single leakage-free video-level split (80%:20%, training:testing), stratified by seizure type and video ID for both modeling approaches.

### Pose Estimation Model

We used an open-source deep learning software (multi-animal DeepLabCut/DLC; Figure 2)^9,17^ to estimate body position by tracking anatomical landmarks across representative video frames. Twenty representative video frames were selected from each event video by k-means clustering to capture diverse postures and movements. After being trained with a labeling protocol, three neurology residents with at least two years of neurology training (SM, MM, IL) used DLC’s annotation interface^18^ to manually label each set of 20 frames with 26 anatomical landmarks. These landmarks were selected by the project team as having the highest value in differentiating seizure etiology (Table S1). We trained the DLC model on the labeled frames to predict 2D coordinates (*keypoints*) of each landmark across all frames. To ensure reproducibility and generalizability, the DLC model was trained in three separate random frame “shuffles” (Data Supplement) with a 95 % training and 5% testing split (Table S2).

**Figure 2.**
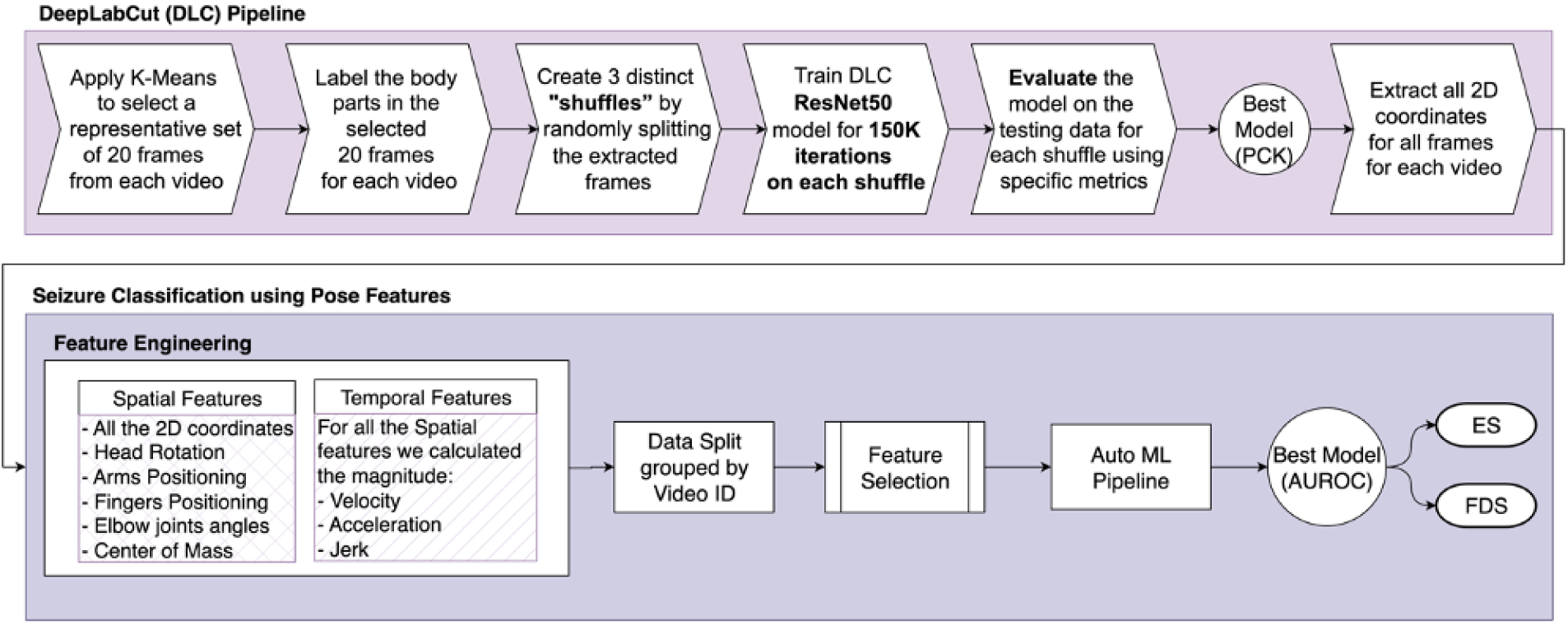
Pose estimation classifier end-to-end pipeline.

Using head size as a scaling threshold (based on prior work), 1/17/2026 12:16:00 AM^19–21^ we calculated several standard metrics to evaluate DLC model performance: (1) *percentage of correct keypoints* (PCK), which defined a “correct” keypoint if the Euclidean distances between predicted and true locations were ≤50% of threshold value; (2) *global localization error,* by root mean squared error (RMSE) normalized to threshold across all landmarks and videos, and (3) sensitivity, accuracy, precision, and area under receiver-operating (AUROC) and precision-recall curves (AUPRC), treating keypoints as “correct” or “incorrect” based on the scaling threshold (Data Supplement). Because many patients were monitored while supine and/or under bedsheets, we also introduced (4) *occlusion detection* to assess the model’s ability to identify occluded landmarks^22,23^. Prior to using the DLC model landmark predictions for our pose-estimation-based classifier, we evaluated the performance of DLC model landmark predictions using PCK across different scaling thresholds.

After training and testing the DLC model, we used the model with the highest PCK on both training and testing to predict the position of all keypoints across all frames in each video, thereby generating a time-resolved series of 2D coordinates. Using these coordinates, we derived a diverse set of spatial and temporal features that captured body motion patterns with clinical significance, in addition to motion dynamics, such as *velocity*, *acceleration*, and *jerk* (Data Supplement).

We then reduced the dimensionality of all candidate training features by applying two complementary univariate feature selection methods: *analysis of variance* (ANOVA) and *mutual information* (MI). In ANOVA, we selected the 50 highest F1-scoring features among those with statistically significant p-values (p < 0.05). In MI, we used a threshold of 0.06 to retain features from the right tail of the MI-score distribution. In parallel, we applied principal component analysis (PCA) to the entire training feature set. Finally, we combined the unique ANOVA and MI selected features with these PCA components to form our final feature set. We applied this same selection and dimensionality reduction to the testing data.

Using the final feature set, we implemented an existing auto-ML pipeline, (H2O.ai, v.3.46.0.6)^24^ which trained and tuned an ensemble of classification models via 5-fold-cross-validation on the training set, selecting the best-performing model based on AUROC. For the best performing model, we determined accuracy, AUROC, precision, sensitivity, specificity, and F1-score with respect to the recorded gold standard on the held-out test set.

### ResNet18 Model

As a comparison to the pose estimation methodology, we developed an end-to-end deep learning pipeline by adapting a 3D-CNN (ResNet3D-18)^25^ that was pre-trained on video data and designed to capture both spatial and temporal patterns from video frames without requiring manual frame annotation. In this approach, all video clips were preprocessed to match the expected input format of the ResNet3D-18 model (i.e., resizing all video clip frames to 128 x 171 pixels, centrally cropping all video frames to 112 x 112 pixels, and applying normalization parameters from the original training dataset). To improve generalizability, we also applied data augmentation techniques during training (Data Supplement).

We employed 3-fold cross-validation within the training set, training each fold with an Adam optimizer and cross-entropy loss, and saving the model checkpoint yielding the lowest validation loss. After cross-validation, the best-performing model was retrained on the entire set and evaluated on a held-out test set to assess generalizability. The held-out test set was identical to that used to evaluate the DLC-autoML pipeline, allowing for direct performance comparisons between both approaches. Classification performance was assessed using accuracy, AUROC, AUPRC, precision, sensitivity, specificity, and F1-score.

### Statistical Analysis

Categorical variables were summarized as counts and percentages by seizure type and compared using Pearson’s chi-square or Fisher’s exact tests wherever appropriate. Continuous measures that deviated from normality were reported as median and interquartile range (IQR) and compared between groups using two-sided Mann–Whitney U test. To control the family-wise error rate arising from multiple comparisons, raw p-values were adjusted with the Bonferroni method. Alpha was set at 0.05. All analyses were performed on complete-case data for each variable, and statistical computations were carried out using SciPy v.1.12.0. All models were run in Python v.3.12.2 on NVIDIA V100 GPUs using our institution’s high-performance computing environment^26^.

For the overall best-performing model, we evaluated six probability cutoffs—0.10, 0.25, the threshold maximizing Youden’s J statistic^27^ (sensitivity + specificity − 1), 0.50 (default), 0.75, and 0.90 to identify an optimal decision boundary for inference. Youden’s J threshold was determined on the validation set and subsequently applied to the held-out test set.

## RESULTS

Over the study period, we extracted 51 FDS and 70 ES video clips, of which 6 FDS and 9 ES events were excluded due to low video quality or mostly occluded frames. In our final analysis, we included 106 event videos (2,097 frames) comprising 61 (57.5%) ES and 45 (42.5%) FDS videos and corresponding to 101 unique patients. Median video length was 5.2 (IQR: 1.0) minutes, with FDS event videos being significantly longer than ES videos (median (IQ) duration 5.75 (2.32) vs. 5.15 (0.6) minutes, p = 0.03) (Table S3).

Of the 101 patients, the median cohort age was 27.5 (IQR 22.0) years; 59 (58.4%) had at least one ES event (“ES group”) and 42 (41.6%) had at least one FDS event (“FDS group”; Table 1). In comparison to patients in the ES group, patients in the FDS group were significantly more likely to be female (88.1% vs. 37.3%, p < 0.01) and less likely to be White (9.5% vs. 35.6%, p=0.02) and have undergone EEG monitoring in epilepsy monitoring units (16.7% vs. 42.4%, p = 0.04). By contrast, patients in the ES group were significantly more likely to have brain MRI evidence of structural abnormalities (35.7% vs. 74.6%, p = 0.01).

**Table 1.**
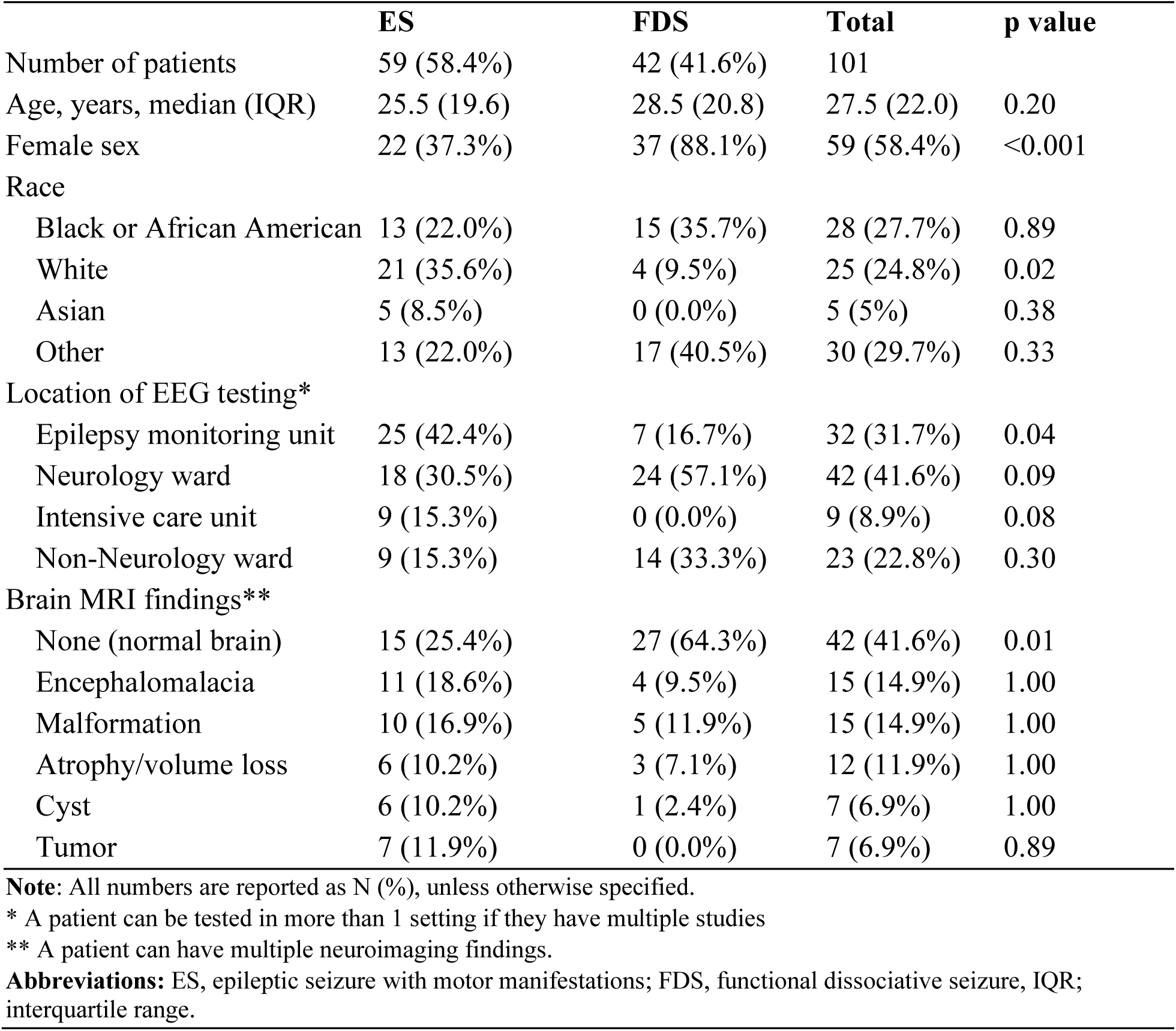
Description of video data set, stratified by event type.

|  | ES | FDS | Total | p value |
| --- | --- | --- | --- | --- |
| Number of patients | 59 (58.4%) | 42 (41.6%) | 101 |  |
| Age, years, median (IQR) | 25.5 (19.6) | 28.5 (20.8) | 27.5 (22.0) | 0.20 |
| Female sex | 22 (37.3%) | 37 (88.1%) | 59 (58.4%) | <0.001 |
| Race |  |  |  |  |
| Black or African American | 13 (22.0%) | 15 (35.7%) | 28 (27.7%) | 0.89 |
| White | 21 (35.6%) | 4 (9.5%) | 25 (24.8%) | 0.02 |
| Asian | 5 (8.5%) | 0 (0.0%) | 5 (5%) | 0.38 |
| Other | 13 (22.0%) | 17 (40.5%) | 30 (29.7%) | 0.33 |
| Location of EEG testing* |  |  |  |  |
| Epilepsy monitoring unit | 25 (42.4%) | 7 (16.7%) | 32 (31.7%) | 0.04 |
| Neurology ward | 18 (30.5%) | 24 (57.1%) | 42 (41.6%) | 0.09 |
| Intensive care unit | 9 (15.3%) | 0 (0.0%) | 9 (8.9%) | 0.08 |
| Non-Neurology ward | 9 (15.3%) | 14 (33.3%) | 23 (22.8%) | 0.30 |
| Brain MRI findings** |  |  |  |  |
| None (normal brain) | 15 (25.4%) | 27 (64.3%) | 42 (41.6%) | 0.01 |
| Encephalomalacia | 11 (18.6%) | 4 (9.5%) | 15 (14.9%) | 1.00 |
| Malformation | 10 (16.9%) | 5 (11.9%) | 15 (14.9%) | 1.00 |
| Atrophy/volume loss | 6 (10.2%) | 3 (7.1%) | 12 (11.9%) | 1.00 |
| Cyst | 6 (10.2%) | 1 (2.4%) | 7 (6.9%) | 1.00 |
| Tumor | 7 (11.9%) | 0 (0.0%) | 7 (6.9%) | 0.89 |

**Note:** All numbers are reported as N (%), unless otherwise specified.
\* A patient can be tested in more than 1 setting if they have multiple studies
\*\* A patient can have multiple neuroimaging findings.
**Abbreviations:** ES, epileptic seizure with motor manifestations; FDS, functional dissociative seizure, IQR; interquartile range.

### Pose Estimation Pipeline Performance

Training and testing PCK values increased with threshold values, plateauing at approximately 40%. Mean PCK across all thresholds was 0.90 (SD ± 0.20) for the training set and 0.87 (SD ± 0.50) for the testing set (Figure S1) with the highest training and testing AUROC-PCK of 0.91 and 0.88, respectively, for Shuffle 1. The DLC model’s overall scaled RMSE was 0.18 (SD ± 0.62) pixels in the training set and 0.25 (SD ± 0.42) pixels in the testing set. Facial landmarks exhibited consistently low RMSE (i.e., high accuracy) with 0.12 (SD ± 0.08) in training and 0.15 (SD ± 0.11) in testing, whereas appendicular landmarks showed greater RMSE values (arm RMSE 0.26, SD ± 0.47 training; 0.36, SD ± 0.62 in testing) (leg RMSE 0.37, SD ± 0.79 for training; 0.40, SD ± 0.63 for testing; Figure S2). Overall, most landmark RMSE values were below the scaling threshold, suggesting reliable spatial localization.

When binarizing the predictions, the model’s AUROC was 0.83 (SD ± 0.01) and 0.80 (SD ± 0.01) in training and testing sets, respectively (Figure S3). Precision was 0.96 (SD ± 0.00) on training and 0.95 (SD ± 0.01) on testing, with sensitivity of 0.75 (SD ± 0.01) on training and 0.74 (SD ± 0.01) on testing. Occlusion prediction achieved sensitivity of 0.95 (SD ± 0.00) in training and 0.94 (SD ± 0.00) in testing whereas precision was 0.88 (SD ± 0.01) in training and 0.82 (SD ± 0.01) in testing (Figure S4).

### Pose-based Seizure Classifier Performance

From the initial 392 features extracted using the Shuffle 1 DLC model, we derived a final set of 60 features for modeling. This included 50 features selected through ANOVA (Table S4), and 30 additional features from MI analysis after accounting for overlap (Figure S5). PCA revealed that the first two components captured the largest portion of shared variance (Figure S6) and were added to complete the final feature set.

The best-performing H2O-auto-ML classifier was a logistic regression model with moderate-penalty ridge (λ=0.5) regularization, achieving a mean cross-validated AUROC of 0.66 (SD ± 0.13). On the held-out test set, the model achieved accuracy 0.62, precision 0.50, sensitivity 0.91, F1 0.65, AUROC 0.71, and AUPRC 0.53. By contrast, ResNet3D-18 had a cross-validated AUROC 0.77 (SD ± 0.12). On the test set using the default probability cut-off, accuracy was 0.80, precision 0.82, sensitivity 0.82, AUROC 0.78, and AUPRC 0.84 (Table 2, Figure 3). The impact of ResNet3D-18 output thresholds on model performance is illustrated in Table 3.

**Table 2:** Classifier Testing Results.

| Table 2: Classifier Testing Results |  |  |  |  |  |  |
| --- | --- | --- | --- | --- | --- | --- |
| Model | Accuracy | Precision | Sensitivity | F-1 Score | AUROC | AUPRC |
| H2O AutoML * | 0.62 | 0.50 | 0.91 | 0.65 | 0.71 | 0.53 |
| ResNet3D-18 | 0.80 | 0.82 | 0.82 | 0.82 | 0.78 | 0.84 |

All model performances are reported using a 0.5 probability threshold.
Abbreviations: AUROC, area under receiver-operating curve; AUPRC, area under precision-recall curve; ML, machine learning.
\* Logistic regression with $\lambda=0.5$ .

**Figure 3.**
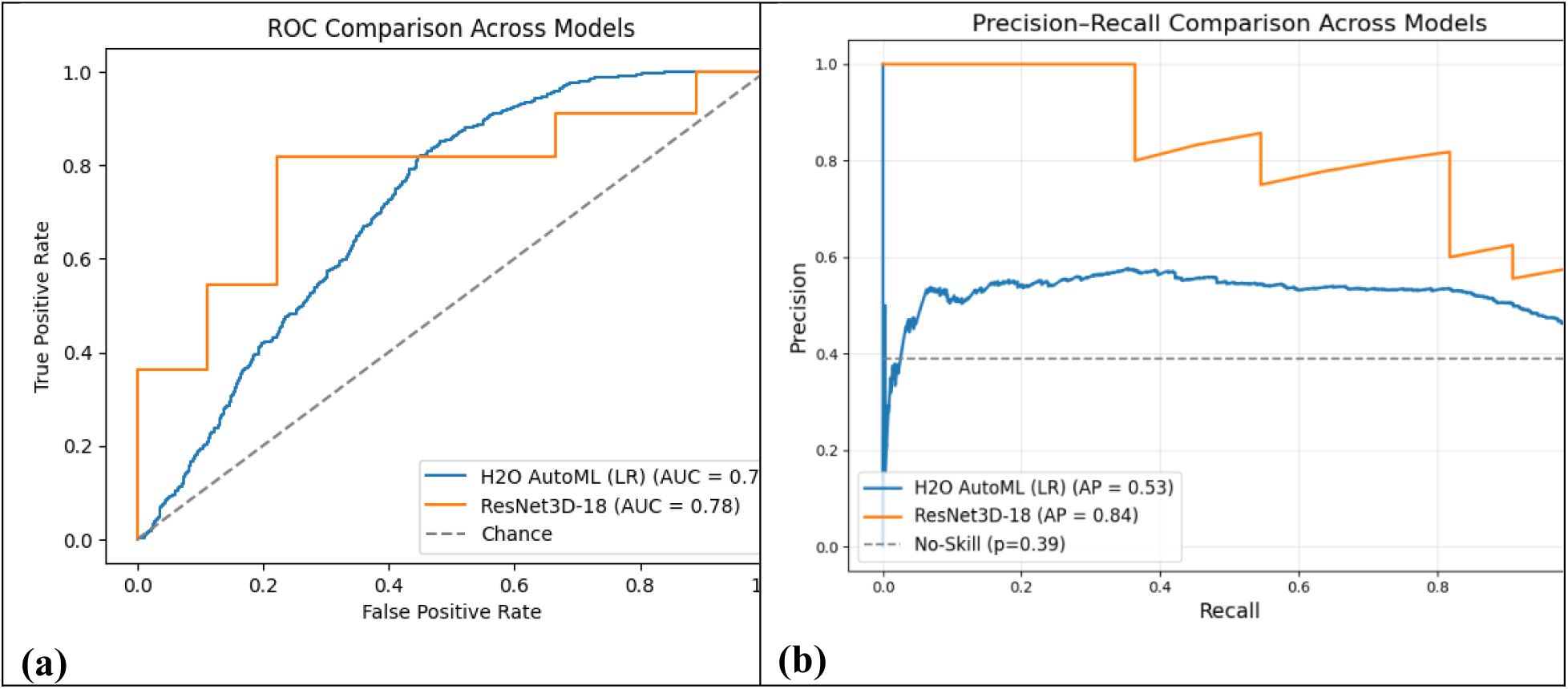
(a) AUROC and (b) AUPRC for the two classifiers on the held-out test set. H2O-Auto-machine learning ridge-penalized logistic regression is illustrated in blue; end-to-end ResNet3D-18 model is illustrated in orange. Dashed gray lines represent random-chance performance (AUROC = 0.50) and the no-skill baseline (AUPRC = 0.39), respectively. Abbreviations: ROC, receiver-operating curve; AUROC, area under receiver-operating curve; AUPRC, area under precision-recall curve.

**Table 3:**
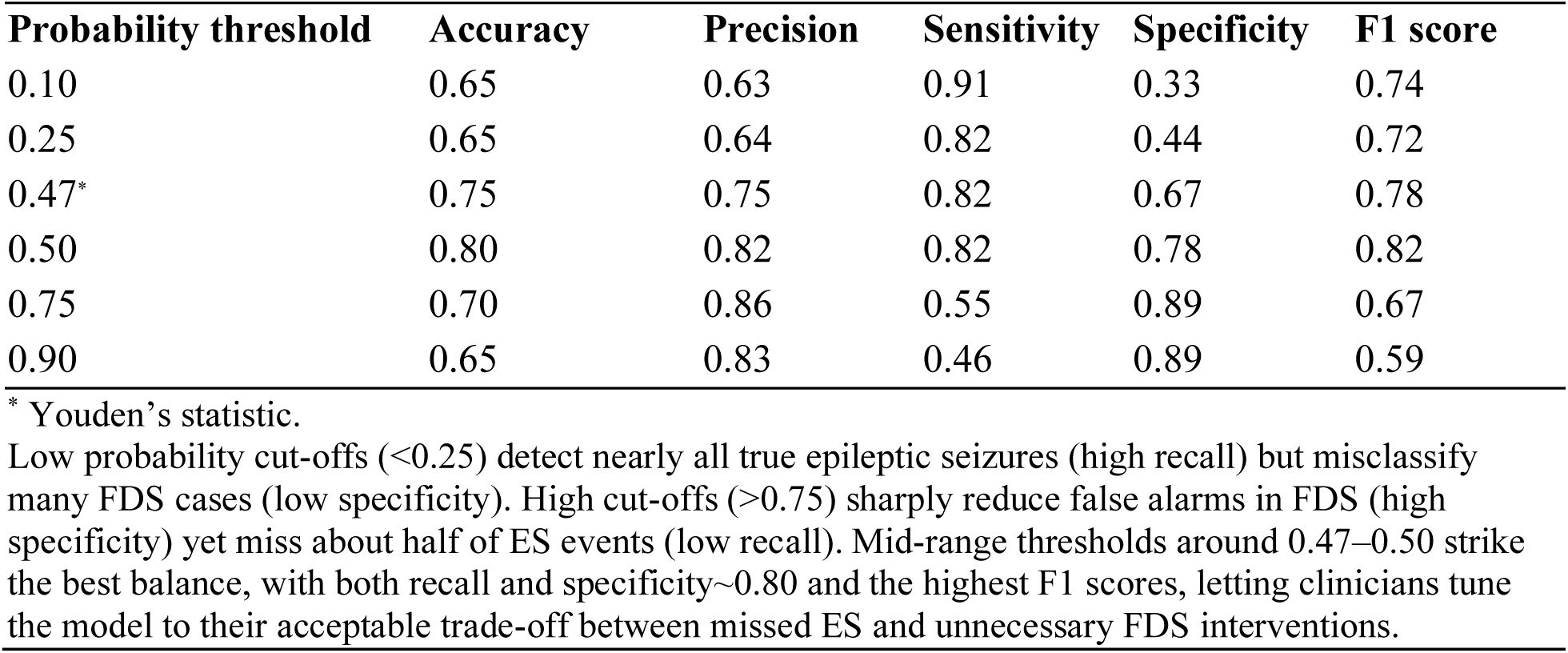
ResNet3D-18 model performance at different probability thresholds.

| Probability threshold | Accuracy | Precision | Sensitivity | Specificity | F1 score |
| --- | --- | --- | --- | --- | --- |
| 0.10 | 0.65 | 0.63 | 0.91 | 0.33 | 0.74 |
| 0.25 | 0.65 | 0.64 | 0.82 | 0.44 | 0.72 |
| 0.47* | 0.75 | 0.75 | 0.82 | 0.67 | 0.78 |
| 0.50 | 0.80 | 0.82 | 0.82 | 0.78 | 0.82 |
| 0.75 | 0.70 | 0.86 | 0.55 | 0.89 | 0.67 |
| 0.90 | 0.65 | 0.83 | 0.46 | 0.89 | 0.59 |

\* Youden’s statistic.
Low probability cut-offs (<0.25) detect nearly all true epileptic seizures (high recall) but misclassify many FDS cases (low specificity). High cut-offs (>0.75) sharply reduce false alarms in FDS (high specificity) yet miss about half of ES events (low recall). Mid-range thresholds around 0.47–0.50 strike the best balance, with both recall and specificity~0.80 and the highest F1 scores, letting clinicians tune the model to their acceptable trade-off between missed ES and unnecessary FDS interventions.

## DISCUSSION

We developed two ML classifiers that distinguish ES and FDS using only videos of events with motor manifestations concerning for seizures. One utilized an intermediary pose estimation model while the other utilized a CNN without pose estimation. The DLC pose estimation model demonstrated high sensitivity in labeling anatomic keypoints, suggesting reliability and consistency in identifying occluded and non-occluded landmarks at our validated scaling threshold with minimal variability across shuffles. While both models demonstrated moderate to good classifying ability, the 3D-CNN approach outperformed the pose estimation approach by all performance metrics (except for sensitivity).

Prior analyses of video-based ML seizure detection and classification systems have shown sensitivities of 75-100% and PPVs of above 85%^28–33^. The sensitivity of both of our deep learning models are within the published ranges found in prior work, although our models’ precision values fall below those of prior analyses. However, it is important to note that few analyses have exclusively sought to develop AI models to distinguish FDS and ES.

These few analyses have notable overlap and differences in comparison to our study. One prior investigation that demonstrated similar model accuracy in distinguishing FDS and convulsive ES^34^ used frequency features derived from temporal variations in light intensity (optical flow oscillations), which we did not use. Another recent analysis of a commercial automated video detection system achieved a remarkably high sensitivity (98%) in distinguishing motor FDS from ES^35^ but only included two FDS cases in its training set, thereby limiting any conclusions about performance.

Our study addresses two gaps in the existing body of work by (a) using deep learning to address the specific diagnostic conundrum of differentiating motor FDS from ES, and (b) comparing pose estimation with pose-agnostic modeling techniques. Within (b), each technique presents significant advantages and drawbacks to be considered for potential implementations.

A key advantage of the 3D-CNN approach is its ability to extract spatiotemporal features directly from raw video, obviating the need for time-consuming manual annotation required in pose-based models^36^. This reduces the time and expertise required for CNN development and removes biases introduced during manual feature selection. Thus, the 3D-CNN model offers significant advantages in settings where rapid scalability and minimal human pre-processing are critical.

By contrast, pose estimation models are more interpretable than CNNs^37^, which is advantageous when dealing with incorrect model predictions. By localizing and quantifying specific body movements, pose models can provide clinicians with transparency into the kinematic differences between FDS and ES events. This can be especially useful in settings where decision-making transparency is essential, such as in medicolegal contexts, discussions with patients and families, or hypothesis-generation for future predictive models.

A prediction that incorrectly identifies an ES event as FDS can lead to severe consequences including SE or sudden death in epilepsy, whereas an incorrectly classified FDS event can result in unnecessary interventions. Despite these points, at default thresholds, our best-performing model had an 18% false positive rate, with an AUPRC suggestive of excellent overall tradeoff between sensitivity and precision. From an operational perspective, these results support the use of this model as a diagnostic adjunct rather than a definitive diagnostic tool.

This study was limited by small sample size. Despite the high volume of vEEG performed at our institution, few vEEG captured clinical events meeting inclusion criteria. While a limitation, this is consistent with prior work, underlining how common it is to not have events captured on vEEG monitoring^5^. Further, we did not use standardized heuristics to identify anatomic keypoints for labeling in our pose estimation model. This resulted in significant variability in the positioning of the keypoint placement across multiple labelers. However, our 3D-CNN approach incorporated video information independently of pose, thereby circumventing this limitation.

## CONCLUSIONS

We developed two ML models capable of distinguishing FDS and ES events using videos alone. This lays the groundwork for future studies that apply advanced data science methods to videos to address this common diagnostic dilemma. The models we developed are scalable and offer significant potential for enhancing diagnostic workflows, especially in resource-limited settings where access to EEG and neurologists is scarce. Future work will seek to replicate these results with larger sample sizes, different video types, and alternative modeling, with the end-goal of accelerating diagnostic precision and care delivery for seizure-like events.

## Data Availability

The datasets generated and/or analyzed during the current study are not publicly available due to challenges in de-identifying patient videos.

https://github.com/asalaerekat/epilepsy-project

## Author Contributions

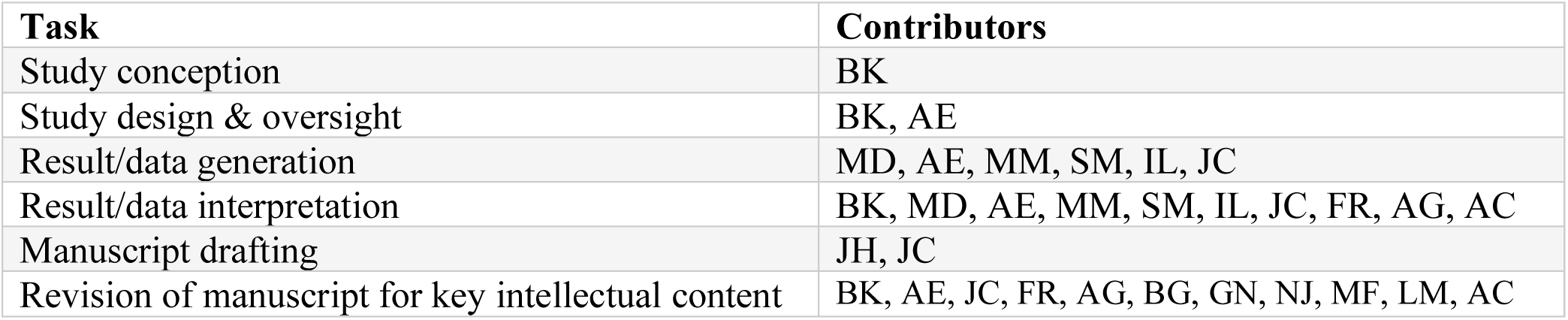

All authors had full access to the data underlying this study and were provided with the opportunity to review this manuscript as well as make appropriate edits.

