## Supplement Text for "A Comparison of Two Deep Learning Approaches to Distinguish Functional Dissociative from Epileptic Seizures Using Event Videos" (from another version of this preprint: version 1, 2025-12-19)

### DATA SUPPLEMENT

#### Video Frame Selection and Keypoint Annotation

We used K-means clustering to generate a diverse and representative training set for pose estimation. This consisted of three steps: (a) feature encoding, in which frames were encoded as high-dimensional feature vectors capturing visual characteristics, (b) K-means clustering to partition frames into 20 clusters based on visual similarity, (c) sampling of one frame per cluster centroid to maximize visual diversity, particularly with respect to body posture and movement states. We chose this unsupervised sampling strategy because it offered improved coverage of motion variability compared to uniform sampling or manual selection, while remaining computationally tractable for large datasets. In preparation for clustering, we defined a total of 26 anatomical landmarks (Supplement Table 1). Three neurology residents with at least two years of neurology experience manually annotated the landmarks using the DeepLabCut (DLC) GUI. Annotation consistency was ensured through a manual review process.

#### Pose Model Training

We trained the pose-estimation model using *multi-animal DeepLabCut (maDLC)* instead of single-animal DLC. Because maDLC produces both heatmaps for landmark detection and part affinity fields (PAFs), which encode the relationships between body parts. During inference, maDLC uses these PAFs to group keypoints into distinct skeletons, ensuring that each set of landmarks belongs to the correct individual. This makes maDLC more robust in situations where multiple subjects, partial occlusions, or overlapping bodies occur, as it reduces the risk of mis-assigning keypoints and maintains accurate identity tracking across frames

We trained three independent maDLC model initializations (or “shuffles”) to assess reproducibility and generalizability. For each shuffle, 95% of labeled frames were used for training and 5% were reserved for testing. We used a ResNet-50 backbone, a batch size of 16, and real-time image augmentation to improve generalization and robustness of the pose-estimation model. Each model was trained for 150,000 iterations using an Adam optimizer with default DLC hyperparameters.

**Pose Estimation Evaluation**

By default, the DLC model output 3 values for each keypoint in every frame: x-coordinate $\hat{x}_{ij}$, y-coordinate $\hat{y}_{ij}$, and *confidence score* $C_{ij}$(ranging from 0 to 1) reflecting the model's certainty that the keypoint is correctly identified. To exclude noisy or unreliable predictions while minimizing data loss, we filtered out low-confidence predictions using a threshold of 0.6. To assess the performance of the maDLC model, we used different metrics derived from pixel-wise localization error between predicted and ground-truth keypoint positions. As a foundation for these metrics, the Euclidean distance between predicted $(\hat{x}_{ij}, \hat{y}_{ij})$ and ground truth $(x_{ij}, y_{ij})$ coordinates was calculated for each keypoint $j$ in frame $i$ as follows:

$$distance_{ij} = \sqrt{(\hat{x}_{ij} - x_{ij})^2 + (\hat{y}_{ij} - y_{ij})^2}\ .$$

From this Euclidean distance, we determined the percent of correct keypoints (PCK), which measures the proportion of keypoints that are correctly localized within a specified threshold. A prediction is considered 'correct' if its Euclidean distance is less than or equal to 50% of the subject's head size. We used head size as a frame-specific normalization factor based on previously published approaches. PCK is expressed as:

$$PCK = \frac{\sum_{i=1}^{N_{\text{frames}}} \sum_{j=1}^{N_{\text{keypoints}}} 1\left(\text{distance}_{ij} \leq 0.5 * headsize_i\right)}{N_{\text{frames}} N_{\text{keypoints}}} \times 100$$

The PCK calculation does not require filtering the keypoints based on the $C_{ij}$ and therefore can determine the overall performance of the model irrespective of the confidence level.

We calculated additional performance metrics as follows:

1. Root mean square error (RMSE): quantifies the average magnitude of keypoint localization error, normalized to the subject's head size to account for scale variability across frames. For each keypoint $j$, RMSE is computed over all frames where the model's prediction confidence $C_{ij}$ exceeds 0.6.

$$RMSE_j = \frac{distance_{ij}}{0.5 * \ headsize_i} \ for \ \forall \ frames \ \{i = 1, \ 2, \ .., \ n\}$$

2. Binary accuracy measures: we evaluated the model's ability to distinguish between coordinate predictions within ('True') or outside ('False') the scaling threshold as a binary task after filtering the keypoints based on the $C_{ij}$. We used accuracy, sensitivity, PPV, and AUC-ROC.

Another important aspect of model performance is its ability to handle occluded keypoints, i.e., keypoints that were not annotated in the video clips due to being blocked by sheets, pillows, or the subject's body position. To assess this, we evaluated whether the model could effectively differentiate between visible and occluded keypoints based on its confidence scores, as follows:

$$O_{ij} = \begin{cases} 1 & \text{if the keypoint is predicted as 'not detected'} \\ 1 - C_{ij} & \text{otherwise} \end{cases}$$

**Feature Engineering from Pose Data**

After reviewing the performance of the DLC model, we used the model with the highest PCK it to generate the x-coordinates, y-coordinates, and confidence scores for each frame across all videos. These coordinates were then used to engineer spatial and temporal features.

*Video and Frame Preprocessing*

To focus on FDS and ES events, each video was trimmed to retain only the clinically relevant segment: the event onset and offset. Based on the recording protocol (2 minutes pre-event and 2 minutes post-event), the first 1 minute and 58 seconds and the final 2 minutes were removed to isolate the event period. Frames with fewer than 25% of visible or non-occluded landmarks were excluded to ensure data quality.

*Spatial Feature Extraction*

Keypoint coordinates were used to compute the following spatial features:

1. Head Rotation: The Euclidean distance between the chin and the midpoint of the left and right shoulders was calculated to quantify head movement relative to the torso.
2. Eye-Chin Distance: The Euclidean distances between the left and right eyes and the chin were computed to capture vertical head positioning.
3. Arm Positioning: For each arm, the average position of the shoulder, elbow, and wrist coordinates was derived to approximate arm posture.
4. Finger Positioning: The average position of all finger keypoints (thumb, index, middle, ring, and pinky) was calculated for each hand. If both hands were tracked, a combined average of both hands was used.

5. Body Center of Mass: The mean of all keypoint coordinates (excluding confidence scores) was computed to estimate the body's spatial center.
6. Elbow Joint Angles: The angle at the elbow joint (Shoulder-Elbow-Wrist) was calculated using the law of cosines. Angles were classified into five categories: "Straight Arm," "Slightly Bent," "Moderately Bent," "Highly Bent," or "Folded/Closed Position."

*Temporal Feature Extraction*

Temporal derivatives are used to capture distinct aspects of movement dynamics:

1. Velocity: First-order frame-wise position change, scaled to units/second (25 fps). This quantifies the speed and direction of a limb or body segment movement, which can help in distinguishing between gradual and rhythmic movements as opposed to rapid and forceful movements, for example. Velocity is expressed as

$$v(t) = \frac{\Delta Position}{\Delta t} = \frac{p(t) - p(t-1)}{1/fps}$$

   where p(t) is the keypoint coordinate at frame t.

2. Acceleration: Second-order derivative of position change, reflecting changes in velocity. This metric highlights the start and stop of a movement and is expressed as

$$a(t) = \frac{\Delta v}{\Delta t} = \frac{v(t) - v(t-1)}{1/fps}$$

3. Jerk: Third-order derivative of position change, capturing abrupt motion changes and measuring the rate of change of acceleration and quantifying 'smoothness' of movement. Higher jerk values signal more abrupt movements. Jerk is expressed as

$$j(t) = \frac{\Delta a}{\Delta t} = \frac{a(t) - a(t-1)}{1/fps}$$

To reduce dimensionality, the magnitude of motion for each keypoint was derived as the Euclidean norm of its x and y velocity, acceleration, and jerk components.

$v_{mag} = \sqrt{v_x^2 + v_y^2}$, $a_{mag} = \sqrt{a_x^2 + a_y^2}$, $j_{mag} = \sqrt{j_x^2 + j_y^2}$, where $v$, $a$, and $j$ denote velocity, acceleration, and jerk respectively.

*Post Feature Engineering Processing*

To ensure robust analysis, we implemented a structured pipeline for handling data quality, normalization, and dimensionality reduction:

1. Missing Data Handling
    a. Threshold Filtering: Intervals with >50% missing features were excluded to eliminate low-quality data.
    b. Temporal Interpolation: Remaining missing values were linearly interpolated using adjacent intervals from the same video, leveraging temporal continuity in motion patterns.
    c. Multivariate Imputation: Persistent gaps were imputed using an iterative chained-equation approach (ICE), which models feature correlations to estimate plausible values.
2. Feature Normalization and Encoding
    - Continuous Features: Standardized to zero mean and unit variance to equalize feature scales.
    - Categorical Variables: Elbow-angle states (e.g., "Straight Arm", "Highly Bent") were converted into binary vectors via one-hot encoding.
3. Dimensionality Reduction

    Principal Component Analysis (PCA) was applied to capture dominant motion patterns across features. The first two principal components, explaining the largest variance, were appended to the original feature set. This hybrid approach retained interpretability of raw features while enriching the model's capacity to identify latent motion synergies.

#### Classification Framework

We designed a classification pipeline with rigorous validation and automated model selection. We split the data into 80% training and 20% testing with 5-fold cross validation for hyperparameter tuning within the training set. To prevent any data leakage, we partitioned the frames within each split by the video level so any set of frames from the same video were ensured to be in the same split/fold. We deployed H2O AutoML to systematically compare different machine learning models or an ensemble of these models, including gradient-boosted machines (GBM), deep neural networks (DNNs), and stacked ensembles. The framework optimized for area under the precision-recall curved (AURPC) while enforcing early stopping to prevent overfitting. We evaluated the performance using Accuracy, AUCROC, PPV, Sensitivity, and F1-score.

### VIDEO CLASSIFICATION PIPELINE: 3D ResNet-18 Model

#### Video Preprocessing

In parallel with the pose estimation approach, we also explored the use of a deep learning model, specifically a 3D ResNet-18, to directly classify seizures from raw video data. Like the pose estimation preprocessing, each video was trimmed to exclude the first 1 minute and 58 seconds and the final 2 minutes, focusing on the segments of the video that contain clinically relevant seizure activity. The spatial resolution of the video was resized to 112 by 112 pixels, but we kept the original 25 fps. Additionally, grayscale videos were converted to RGB format, ensuring consistency in input format.

To augment the robustness of the classifier and prevent overfitting, each video underwent additional data augmentation strategies, including random rotations up to 10 degrees and slight alterations in brightness, contrast, saturation, and hue. This augmentation diversified the training set, enhancing the generalizability of the model to variations commonly observed in clinical practice.

**3D ResNet-18 Model architecture**

The 3D ResNet-18 model was employed to classify seizures directly from the raw video data, leveraging the spatial and temporal dimensions of the video. Unlike the pose estimation method, which extracts key body landmarks, the 3D ResNet-18 model operates on the full video input, learning complex spatiotemporal features that capture the dynamics of seizure-like events. The model's architecture consists of residual blocks that enable it to effectively process both spatial and temporal features, making it well-suited for video data where both dimensions are critical for accurate classification.

**Model training and evaluation**

We partitioned the dataset into training, validation, and testing sets, with an 80/20 split between training/validation and testing data. To ensure robustness and minimize bias, we employed 3-fold cross-validation on the training and validation sets. During each fold, the model was trained using the Adam optimizer and a cross-entropy loss function, with performance monitored using metrics such as accuracy, precision, recall, specificity, F1-score, and AUC-ROC. At the end of each fold, the model with the lowest validation loss was retained for further evaluation. Post-validation, we evaluated the model on a held-out testing set to assess its generalizability and clinical applicability.

### COMPUTATIONAL RESOURCES

All training was conducted on the Minerva high-performance computing cluster at Mount Sinai, utilizing two NVIDIA V100 GPUs to enable efficient processing and parallelization of video-based deep learning models.

### SUPPLEMENTAL TABLES

| Table S1: Anatomical Landmarks for DLC Pose Estimation Learning | |
|---|---|
| (1) Left Eye | (14) Right Eye |
| (2) Nose | (15) Chin |
| (3) Left Shoulder | (16) Right Shoulder |
| (4) Left Elbow | (17) Right Elbow |
| (5) Left Wrist | (18) Right Wrist |
| (6) Left Thumb | (19) Right Thumb |
| (7) Left Index Finger | (20) Right Index Finger |
| (8) Left Middle Finger | (21) Right Middle Finger |
| (9) Left Ring Finger | (22) Right Ring Finger |
| (10) Left Pinky | (23) Right Pinky |
| (11) Left Hip | (24) Right Hip |
| (12) Left Knee | (25) Right Knee |
| (13) Left Ankle | (26) Right Ankle |

**Table S2: Dataset partitioning and leakage-prevention strategy for pose-estimation and end-to-end video classifiers**

| **Pipeline stage** | **Objective** | **Unit of split** | **Split ratio** | **Additional safeguards** |
|---|---|---|---|---|
| **Pose-estimation network (DLC)** | Learn 2-D keypoint coordinates | Annotated frames | 95 % train / 5 % internal test | Three random "shuffles" to assess reproducibility; no seizure labels used at this stage |
| **Pose-based classifier** | Distinguish FDS vs ES from keypoint-derived features | Entire videos | 80 % train / 20 % held-out test | Stratified by seizure type and grouped by video so no frame from a given video leaks across splits |
| **End-to-end video classifier (ResNet-18 3D)** | Distinguish FDS vs ES directly from raw video | Entire videos | 80 % train / 20 % held-out test | Identical patient/video cohorts as the pose-based classifier, preserving the leakage-free grouping |

**Abbreviations:** DLC, DeepLabCut; FDS, functional dissociative seizure; ES, epileptic seizure

**Table S3: Summary of Video Data, stratified by event type ***

| | **Total** | **ES** | **FDS** |
|---|---|---|---|
| **Event videos** | 106 | 61 (57.5%) | 45 (42.5%) |
| **Extracted frames** | 2,097 | 1,207 (57.6%) | 890 (42.4%) |
| **Videos per patient, median (IQR)** | 1.0 (0.0) | 1.0 (0.0) | 1.0 (0.0) |
| **Video length, minutes, median (IQR)** | 5.18 (1.04) | 5.15 (0.6) | 5.75 (2.32) |

*Initial videos identified were 121; all numbers are reported as N **(**%) unless otherwise noted.
Abbreviations: ES, epileptic seizure; FDS, functional dissociative seizure; IQR, interquartile range.

**Table S4. Top 50 DLC-derived pose features ranked by one-way ANOVA F-statistic significance.**

| DLC-based Feature | F-statistic | p_value |
|---|---|---|
| **RightHip_y_std** | 522.6849 | 3.7E-112 |
| **left_arm_x_mean** | 484.1828 | 2.9E-104 |
| **Velocity_mag_RightHip_std** | 437.4638 | 1.19E-94 |
| **acceleration_mag _RightHip_std** | 403.2781 | 1.41E-87 |
| **jerk_mag _RightHip_std** | 394.6322 | 8.79E-86 |
| **LeftHip_x_mean** | 377.4365 | 3.29E-82 |
| **RightHip_x_std** | 373.9535 | 1.74E-81 |
| **RightKnee_y_mean** | 368.7157 | 2.15E-80 |
| **left_arm_state_Straight Arm** | 351.5688 | 8.07E-77 |
| **LeftElbow_x_mean** | 349.9714 | 1.74E-76 |
| **LeftHip_x_std** | 348.6339 | 3.31E-76 |
| **LeftWrist_x_mean** | 345.1187 | 1.79E-75 |
| **LeftShoulder_x_mean** | 338.585 | 4.15E-74 |
| **LeftKnee_y_mean** | 314.5673 | 4.43E-69 |
| **mid_shoulder_x_mean** | 312.8726 | 1.01E-68 |
| **velocity_mag _LeftHip_std** | 303.1483 | 1.1E-66 |
| **acceleration_mag _LeftHip_std** | 264.6073 | 1.43E-58 |
| **left_elbow_angle_mean** | 262.7582 | 3.52E-58 |
| **LeftHip_y_std** | 261.7582 | 5.73E-58 |
| **RightMiddleFinger_y_mean** | 253.7253 | 2.85E-56 |
| **right_fingers_avg_y_mean** | 251.8868 | 6.98E-56 |
| **avg_finger_y_mean** | 243.8876 | 3.43E-54 |
| **jerk_mag _LeftHip_std** | 235.9232 | 1.67E-52 |
| **RightIndexFinger_y_mean** | 225.3104 | 2.97E-50 |
| **LeftThumb_x_mean** | 220.5986 | 2.97E-49 |
| **RightShoulder_x_mean** | 215.7306 | 3.21E-48 |
| **RightRingFinger_y_mean** | 211.9617 | 2.03E-47 |
| **LeftRingFinger_y_mean** | 210.4731 | 4.21E-47 |
| **right_elbow_angle_mean** | 206.7551 | 2.6E-46 |
| **LeftEye_x_mean** | 206.4815 | 2.97E-46 |
| **Chin_x_mean** | 184.5644 | 1.39E-41 |
| **RightThumb_y_mean** | 181.5004 | 6.28E-41 |
| **body_center_x_mean** | 179.2952 | 1.86E-40 |
| **LeftMiddleFinger_y_mean** | 178.7318 | 2.45E-40 |
| **RightEye_y_mean** | 154.0579 | 4.68E-35 |
| **RightShoulder_y_mean** | 151.9014 | 1.36E-34 |
| **LeftEye_y_mean** | 151.3097 | 1.82E-34 |
| **mid_shoulder_y_mean** | 150.0757 | 3.35E-34 |
| **velocity_mag _RightShoulder_std** | 148.491 | 7.33E-34 |
| **right_arm_state_Straight Arm** | 147.6823 | 1.09E-33 |

| | | |
|---|---|---|
| **left_fingers_avg_x_mean** | 147.2633 | 1.34E-33 |
| **acceleration_mag _RightShoulder_std** | 144.7938 | 4.56E-33 |
| **jerk_mag _RightShoulder_std** | 143.1202 | 1.04E-32 |
| **LeftPinky_y_mean** | 142.8107 | 1.22E-32 |
| **head_rotation_mean** | 140.8507 | 3.21E-32 |
| **left_fingers_avg_y_mean** | 138.961 | 8.17E-32 |
| **LeftPinky_x_mean** | 138.9179 | 8.35E-32 |
| **Chin_y_mean** | 138.8692 | 8.55E-32 |
| **acceleration_mag _LeftRingFinger_std** | 137.5954 | 1.61E-31 |
| **RightShoulder_x_std** | 135.7857 | 3.94E-31 |

### SUPPLEMENTAL FIGURES

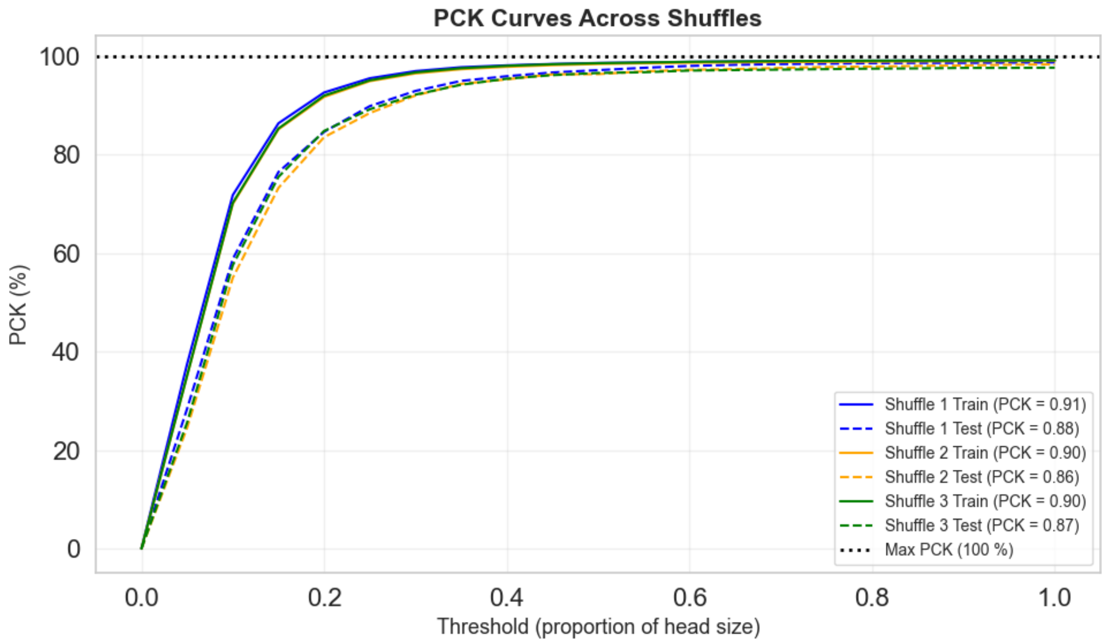


**Figure S1. Percentage of correct keypoints (PCK) in function of scaling threshold.**

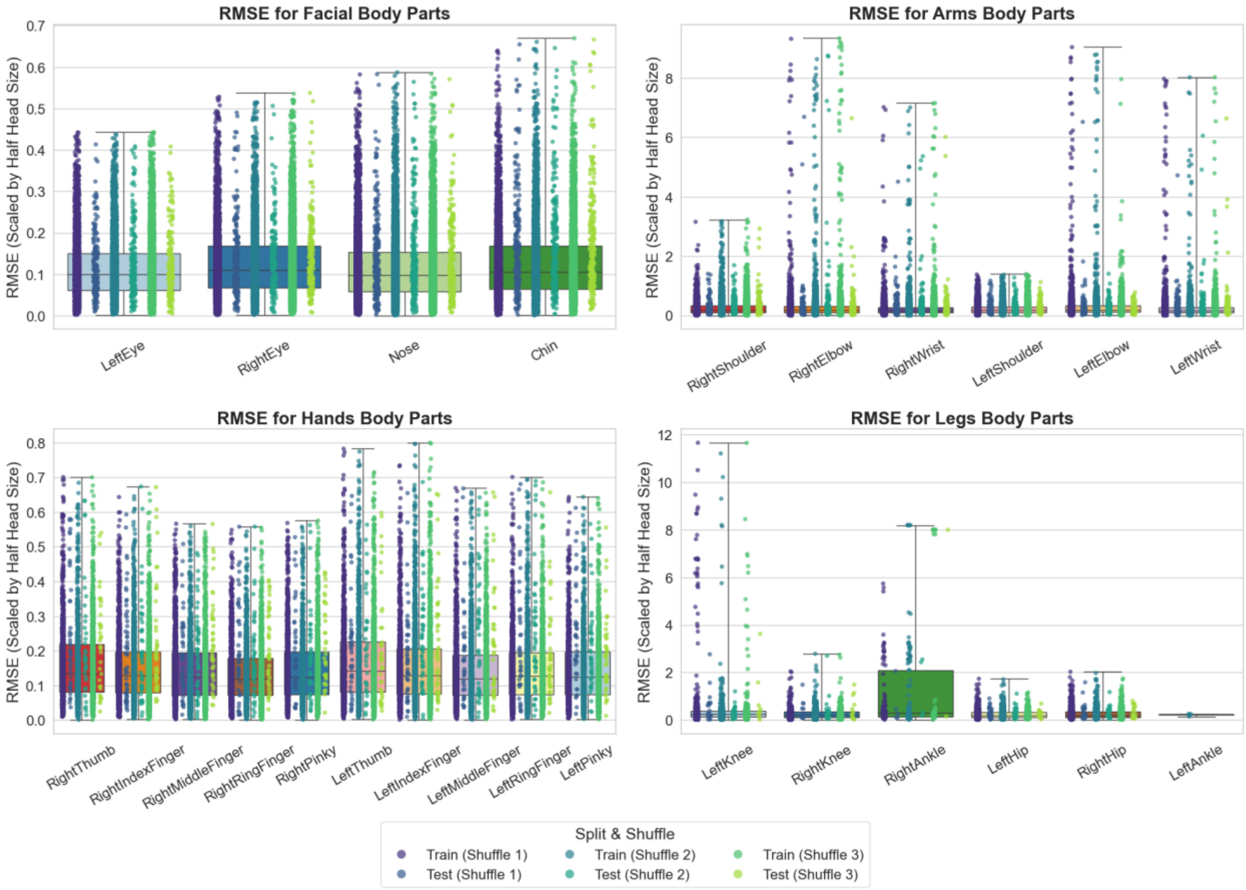


**Figure S2. Root Mean Square Error (RMSE) stratified by labeled anatomic landmark.**

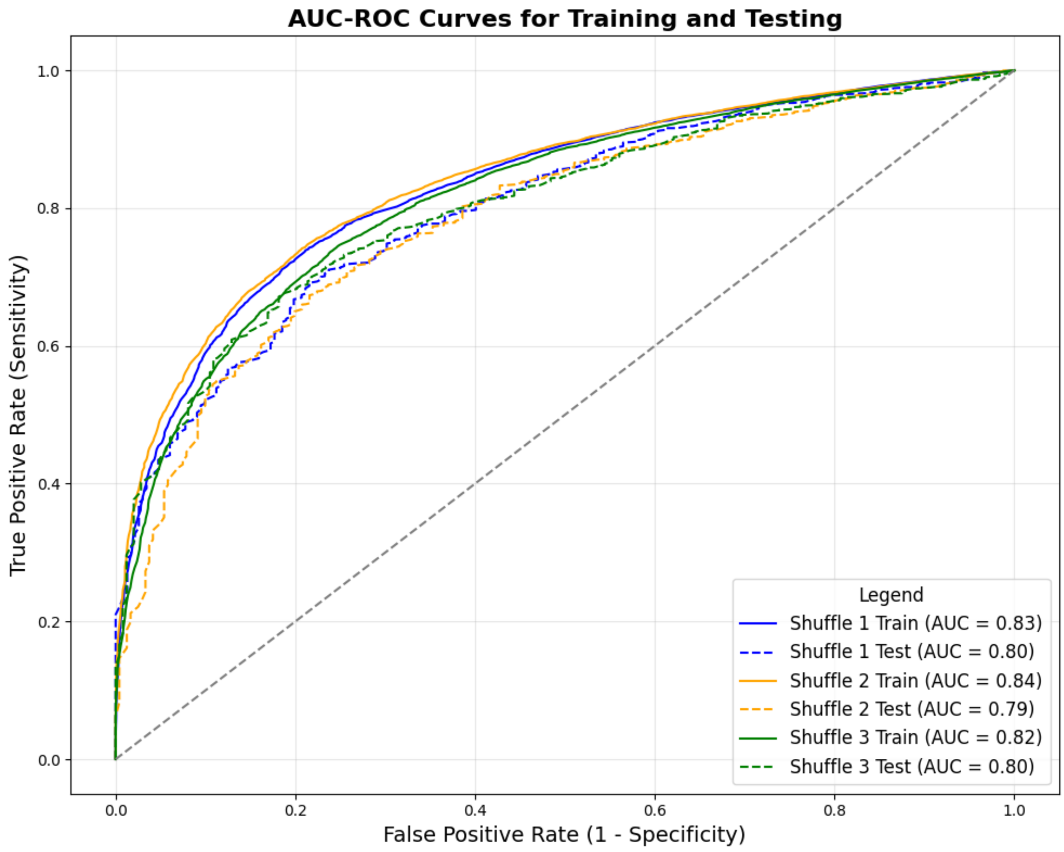


**Figure S3. DeepLabCut pose estimation receiver-operating curve (ROC) across 3 shuffles.**

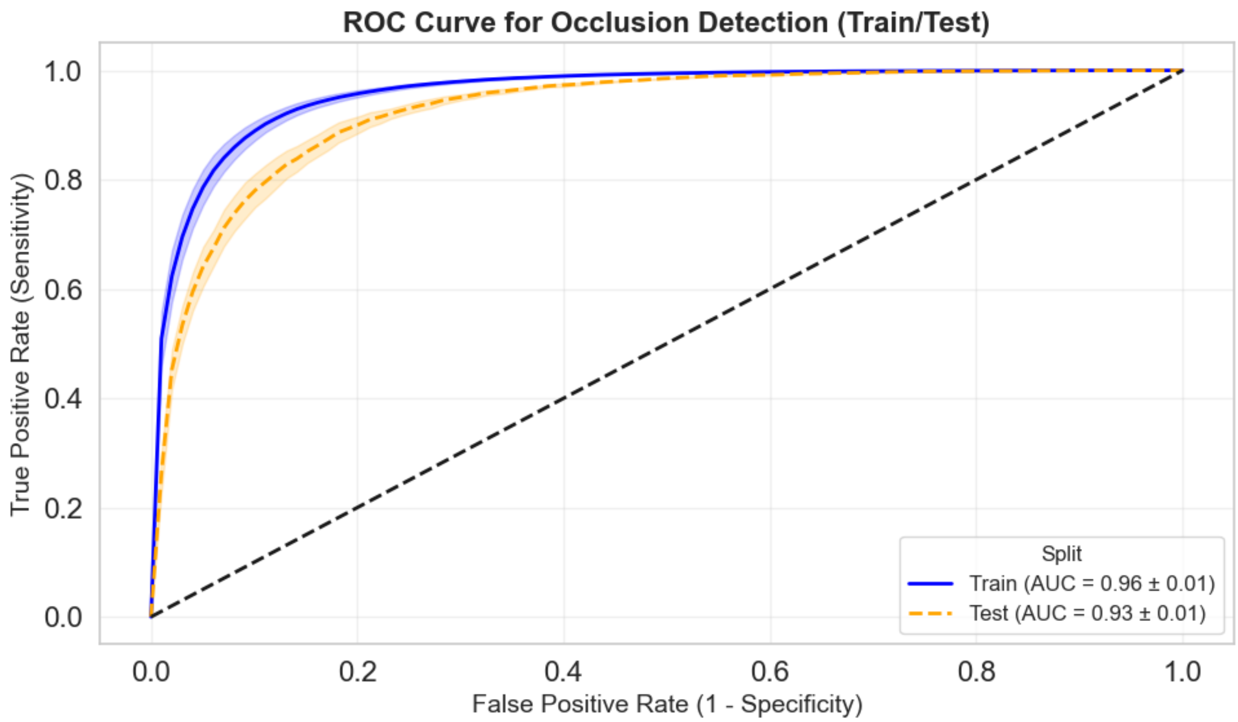


**Figure S4. DeepLabCut pose estimation receiver-operating curve for occlusion detection**

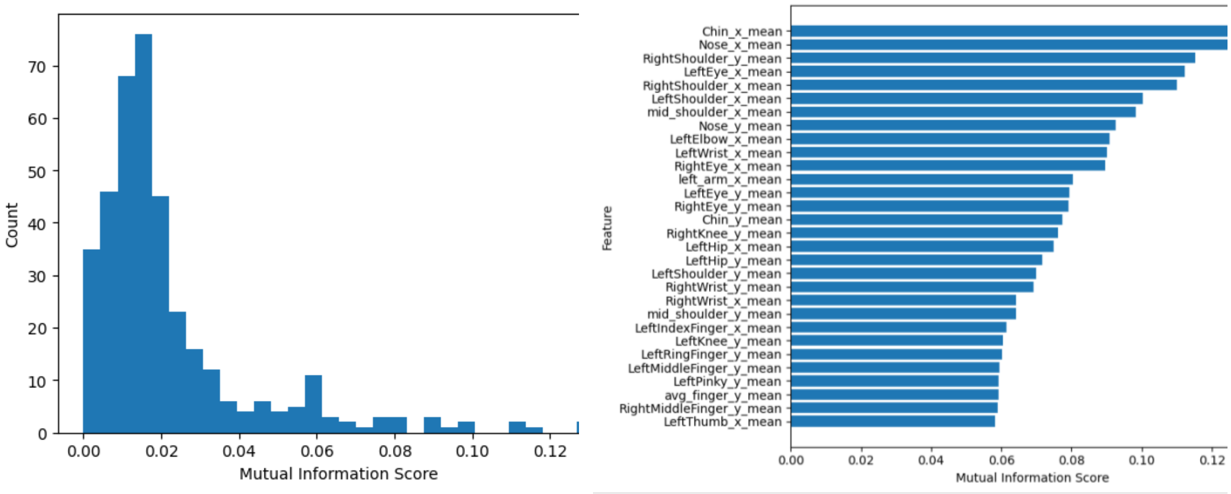


**Figure S5.** (a). Histogram of mutual information scores across DeepLabCut-derived features.

(b). Top 30 DeepLabCut -derived pose features ranked by mutual-information score.

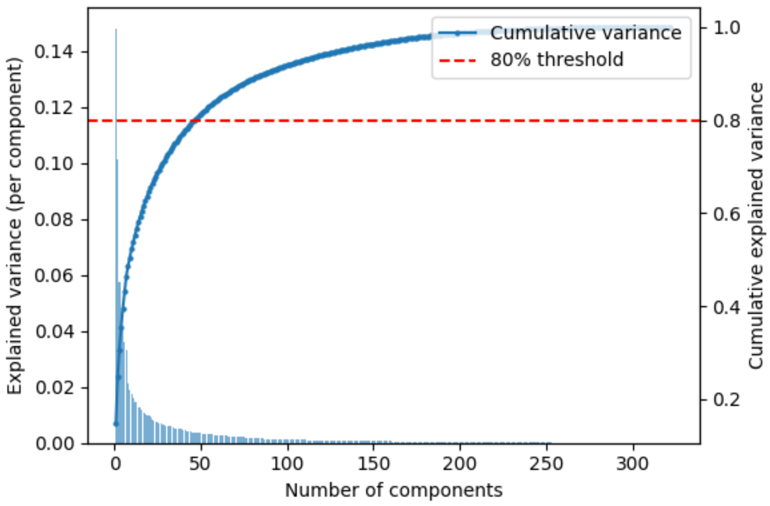


**Figure S6. Scree plot of per-component and cumulative explained variance for principal component analysis on 392 DeepLabCut -derived features.**
